# Combined Effects of Prunus Cerasus (Montmorency Tart Cherry) and Apocynum Venetum (Venetron^®^) On Sleep and Anxiety in Adults with Insomnia

**DOI:** 10.1101/2024.04.24.24306307

**Authors:** Marie Crisel B. Erfe, Paige L. Oliver, Armenouhi Kazaryan, Azure D. Grant, Roy Yoon, Ruchir P. Patel, Belinda Tan, Noah Craft

## Abstract

**Purpose:** Sleep aids derived from traditional plant medicines are strong candidates for safely improving insomnia but require wider validation in patient populations.

**Methods:** We conducted an open label trial of the impact of a compound, Sip2Sleep^**®**^, containing Montmorency tart cherry (*prunus cerasus*) extract and Venetron^**®**^ (*apocynum venetum*) on subjective sleep quality, subjective daytime alertness, sleep duration, sleep latency, anxiety, and insomnia in 43 adults with moderate to severe insomnia. Participants collected data over four weeks, with the sleep aid consumed prior to bed during weeks two and four.

**Results:** The Montmorency tart cherry and Venetron^**®**^ mixture statistically improved subjective sleep quality, daytime alertness, insomnia symptoms, and anxiety without impacting sleep duration and latency. Subjective improvements in sleep quality exhibited a statistical upward trend across the entire study window, suggesting potential persistence of the compounds days after consumption and greater improvement with longer-term consumption.

**Conclusions:** The combination of Montmorency tart cherry and Venetron^**®**^ in this commercially available tincture is a promising sleep aid warranting further investigation in larger trials.

## Introduction

Modern life involves chronic sleep and daily rhythm disruption through artificial light, mistimed meals, and unstable activity schedules; leading to numerous physical^1,2^ and mental maladies^3^. Clinical insomnia may impact 1/5 to 1/3 of the population^4,5^ Additionally, as of 2020, the National Health Interview Survey found that 14.5% of American adults had trouble falling asleep most days or every day in the past month^6^ up from <10% in 1991^7^. Moreover, data on subclinical sleep loss suggests that the majority of Americans are impacted. As of 2013, average sleep duration dropped to 6.8 hours per night, only 56% of people reported getting as much sleep as subjectively needed, and only 34% of Americans report sleeping 8 hours or more per night^8^.

Despite approximately a century of electrified homes in the U.S., today’s widespread use of electronic devices near bedtime, and round-the-clock consumption of stimulating information and food contribute to increasing sleep disruption^9,10^. Such disruption creates a double insult of melatonin suppression via light at night and elevated anxiety related to night-time electronic device usage^11–14^. For example, a recent study of UK adults found light exposure several hours before bed was associated with lower subjective bedtime sleepiness (likely via disruption of rising melatonin^14–16^ and longer sleep onset latency^17^. Similarly, phone usage before bed was associated with poorer sleep quality in two recent Saudi Arabian cohorts^18,19^, and potentially with poorer mood^20,21^. Increasing rates of anxiety and depressive symptoms appear to contribute cyclically to poor sleep quality, with nearly 1/3 American adults reporting anxiety or depressive symptoms in 2023, and nearly 1/2 of youth 18-24^22^. Similar reports of past month anxiety indicate nearly doubling rates in young adults from 2008 to 2018 (from 8% to 14 %), and increases from 6 to 9% in 26-34 year olds^12^.

Together, despite great expansion in the awareness of the importance of sleep^23^, as well as wide uptake in wearable sleep trackers^24,25^, sleep disruption and sleep loss are pervasive. Accordingly, safer interventions that can support quality sleep are needed more than ever. Natural foods and herbal medicines provide a wide array of relatively low-risk potential sleep aids: tart cherry has a long history of safe consumption, while *apocynum venetum* is used in traditional Chinese medicine^26^. This use history provides clues as to safety, preparation, dosage, and potential side effects. The present study investigated the impact of a sleep aid combining two plant extracts traditionally used to improve sleep quality and reduce anxiety: *prunus cerasus* or Montmorency tart cherry, and *apocynum venetum*, Linnaeus (syn. *Trachomitum venetum* L. Woodson) also known as Luobuma in traditional Chinese medicine ^27^ or Venetron^**®**^.

Montmorency tart cherry is the subject of increasing scientific interest due to reported health effects ranging from reducing inflammation and oxidative stress to improving recovery from exercise and boosting sleep quality^28^. Anti-inflammatory and sleep promoting properties are attributed in part to anthocyanins, which may minimize tryptophan degradation while increasing bioavailability for serotonin synthesis. Additionally, sleep-promoting effects arise from large naturally occurring levels of melatonin^28^. Clinical trials and studies on these herbs are limited but suggest potential improvements in sleep quality, duration, and nighttime awakening in athletes and adults with insomnia^29–31^ .

*Apocynum venetum*, derived from the Rafuma leaf, is a lesser-known herb used traditionally in traditional Chinese medicine for calming effects under the name Luobuma^32^. Recent pharmacological studies have explored its antihypertensive effects^32^ and potential as a sleep aid^33^. Active components, including flavonoids and hyperoside, are hypothesized to exert anxiolytic effects in animal models comparable to those of diazepam and buspirone^34^. *Apocynum venetum* is believed to increase GABAergic tone by interacting with GABA receptors, thereby exerting a mild sedating effect, and potentially inducing sleep^34^. To date, however, the authors are aware of only a few studies of *apocynum venetum*’s impact on sleep, reporting improvement in subjective sleep quality and psychological stress^35,36^, alertness^36^, and deep sleep^33^.

To our knowledge, the synergistic effects of these compounds on sleep have not been studied, but the individual components suggest a potential for combined use. Venetron^**®**^’s flavonoids may provide sedative action, whereas melatonin from Montmorency tart cherry may increase sleepiness. This hypothesis aligns with the holistic approach of herbal medicine, where the combined effects of multiple compounds are considered more effective than isolated constituents^37^.Together, Montmorency cherry and Venetron^**®**^ show promise in promoting quality sleep. We hypothesized that their combination in a sleep aid product, called Sip2Sleep^**®**^, would improve subjective and objective sleep parameters and reduce self-assessed insomnia (ISI) and anxiety (GAD-7) in adults with self-reported moderate to severe insomnia.

## Results

*Participatory Research Process and Study Compliance*. Seventy-seven (77) participants were consented and enrolled. Three (3) participants withdrew from the study and twelve (12) were lost to follow-up. Sixty-two (62) participants completed 4 weeks of the study **(Figure 1**) with 98.1% survey compliance in the Chloe (<u>C</u>onsumer <u>H</u>ealth <u>L</u>earning and <u>O</u>rganizing <u>E</u>cosystem) app (**Figure 2**.). In order to be included in the data analysis, participants had to have completed at least 60% of data collection in each arm (Product Use vs. No Product Use), including successfully syncing wearable device data. Forty-three (43) participants met the criteria for final analysis. Devices used were Fitbits (48.8%), Apple Watches (39.5%), Oura Ring (2.3%), Whoop (2.3%), or other devices (4.7%).

**Figure 1.**
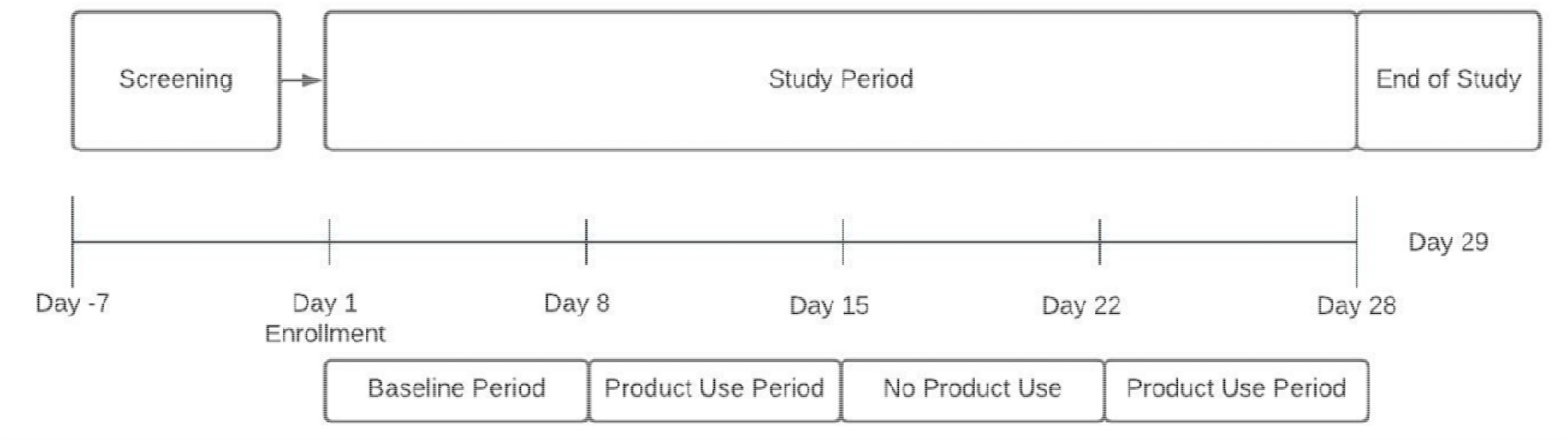
Study Design Timeline. After screening and enrollment, participants completed baseline and daily questionnaires for one week. The sleep aid, Sip2Sleep^**®**^, was taken nightly before bed during weeks 2 and 4, with no intervention taken during week 3. Daily questionnaires and weekly ISI and GAD-7 responses were taken once per week.

**Figure 2.**
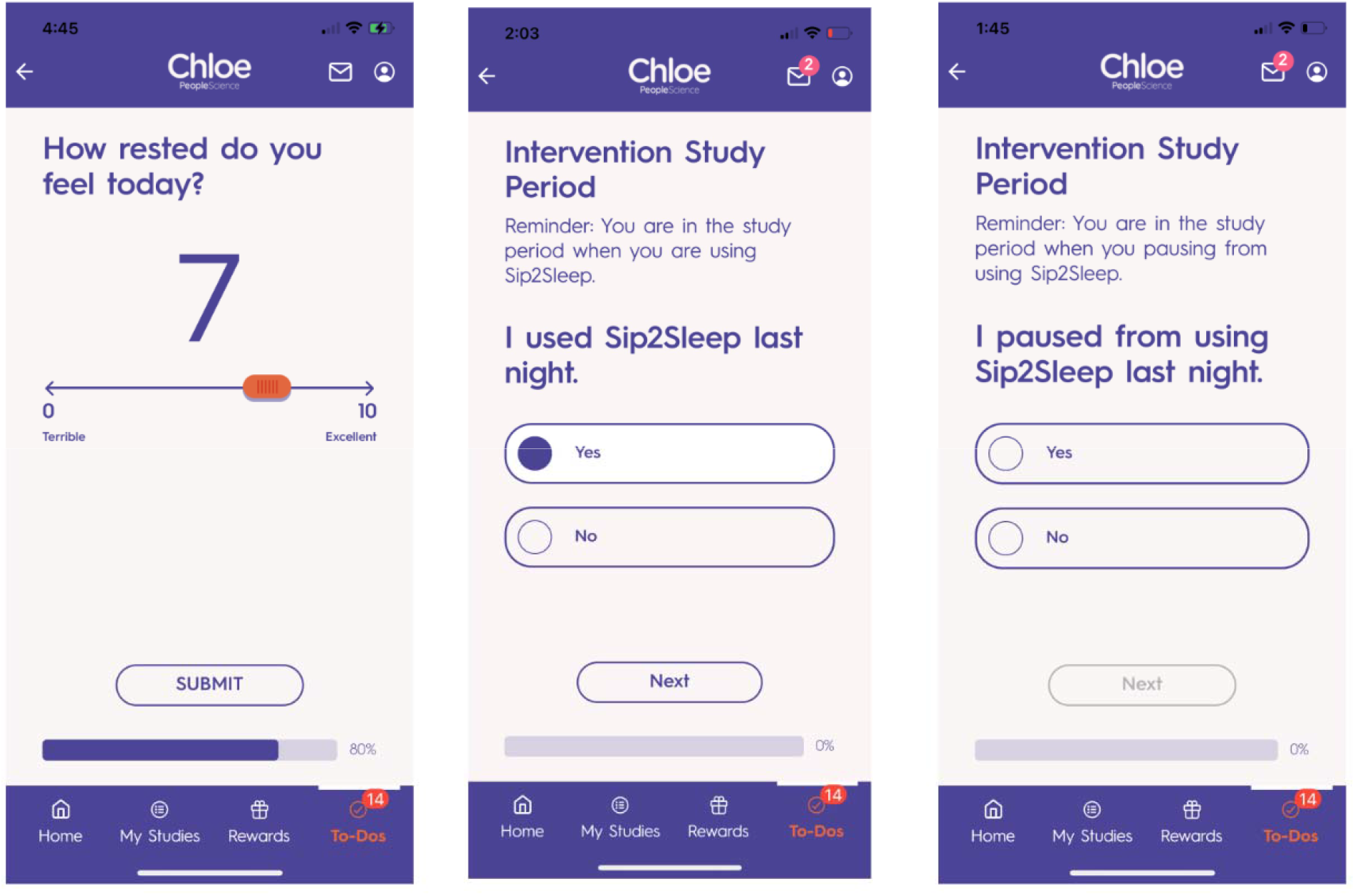
Data Collection in the Chloe Application. **The Chloe App**. Example screens from the Chloe mobile application, where participants gave consent, completed daily questionnaires and health history, received prompts about when to take or omit intervention, and monitored their study progress.

*Demographics*. Participants reported moderate to severe insomnia based on insomnia sleep index (ISI) scores >15 (Morin et al., 2011), (mean = 19.5 +/-3.52) . Mean age was 36.0 +/-12.0 years and the cohort was 75% female. Ethnic backgrounds were as follows: White/Caucasian (86.0%), Black or African American (6.98%), Latino or Hispanic (4.65%), Asian or Asian American (2.32%). Participants reported a range of underlying medical conditions in addition to insomnia, most commonly well-controlled sleep apnea (11.6%), anxiety (6.98%), depression (4.65%), diabetes and heart disease (<3%). Most common medications were Gabapentin (9.3%), Ambien (7.0%), Ozempic (4.7%), progesterone (10% of women), and Prozac (4.7%).

### Sip2Sleep^®^ Impacts Subjective Sleep and Anxiety

Bedtime consumption of Sip2Sleep^**®**^ statistically increased self-reported sleep quality (p=0.003) and daytime alertness (p<0.001), and decreased ISI (p<0.001) and GAD-7 score (p<0.001) **(Figure 3 A-D)**. The compounds did not impact wearable-device generated sleep duration (p=0.64) or latency (p=0.76). **(Figure 3 E,F)**. Results did not differ by sex (data not shown). Additionally, subjective sleep quality and daytime alertness increased statistically week over week during the study (group average increasing Mann-Kendall p<1*10E-4) **(Figure 4A, C)**, whereas sleep duration and latency did not (p=0.19 and p= 0.86, respectively) (**Figure 4 B, D**).

**Figure 3.**
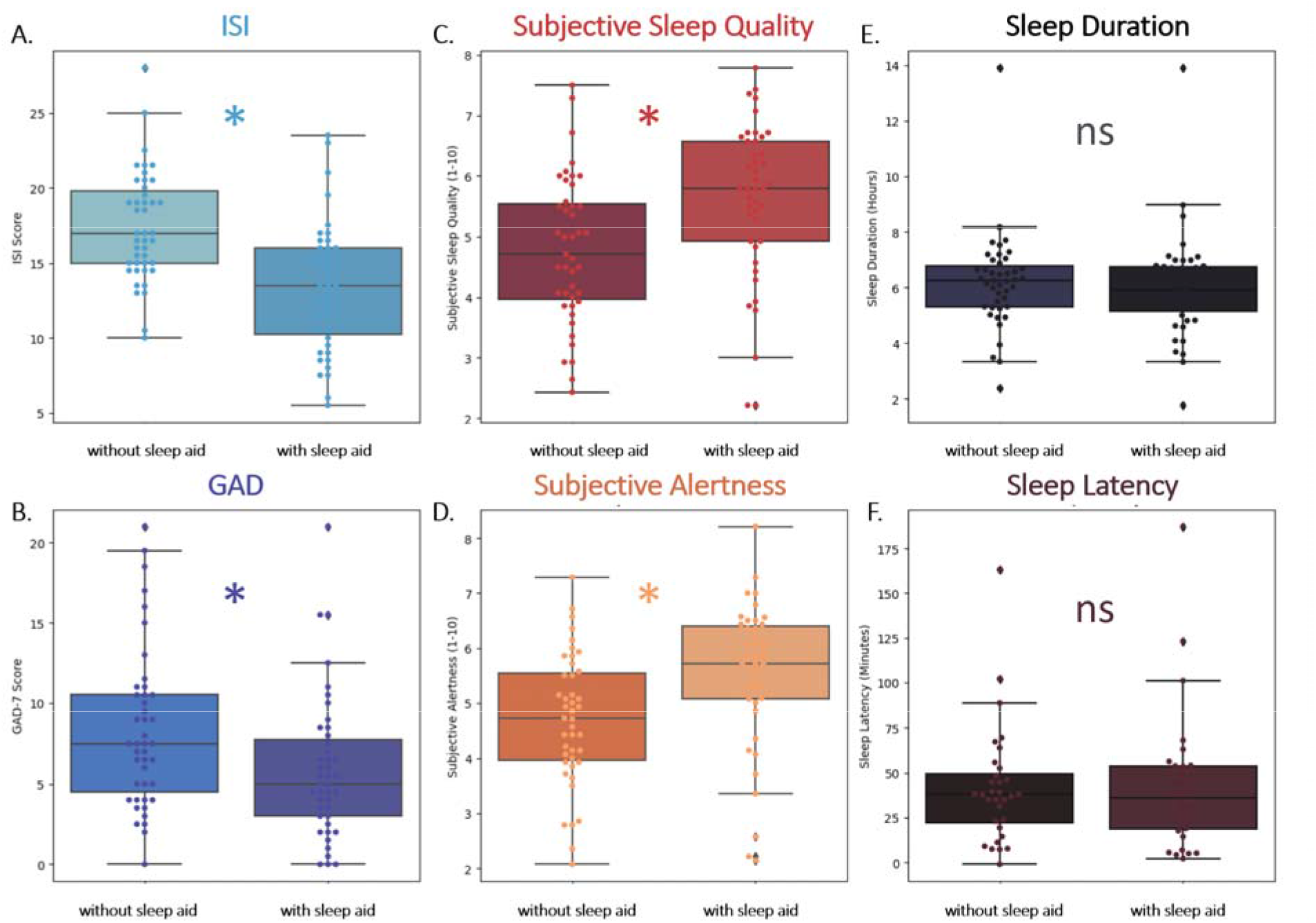
Sip2Sleep^®^ Increases Subjective Sleep Quality, Alertness, and Decrease Insomnia Severity and Anxiety. Box and whisker plots of ISI and GAD-7(A-B, blue), subjective daily sleep quality and alertness (C-D, red and orange), sleep duration and latency (E-F, black and brown) mean scores of all individuals not taking (left side of subplots) and taking (right side of subplots) Sip2Sleep^**®**^. Stars indicate statistical difference (p<0.05) for each metric.

**Figure 4.**
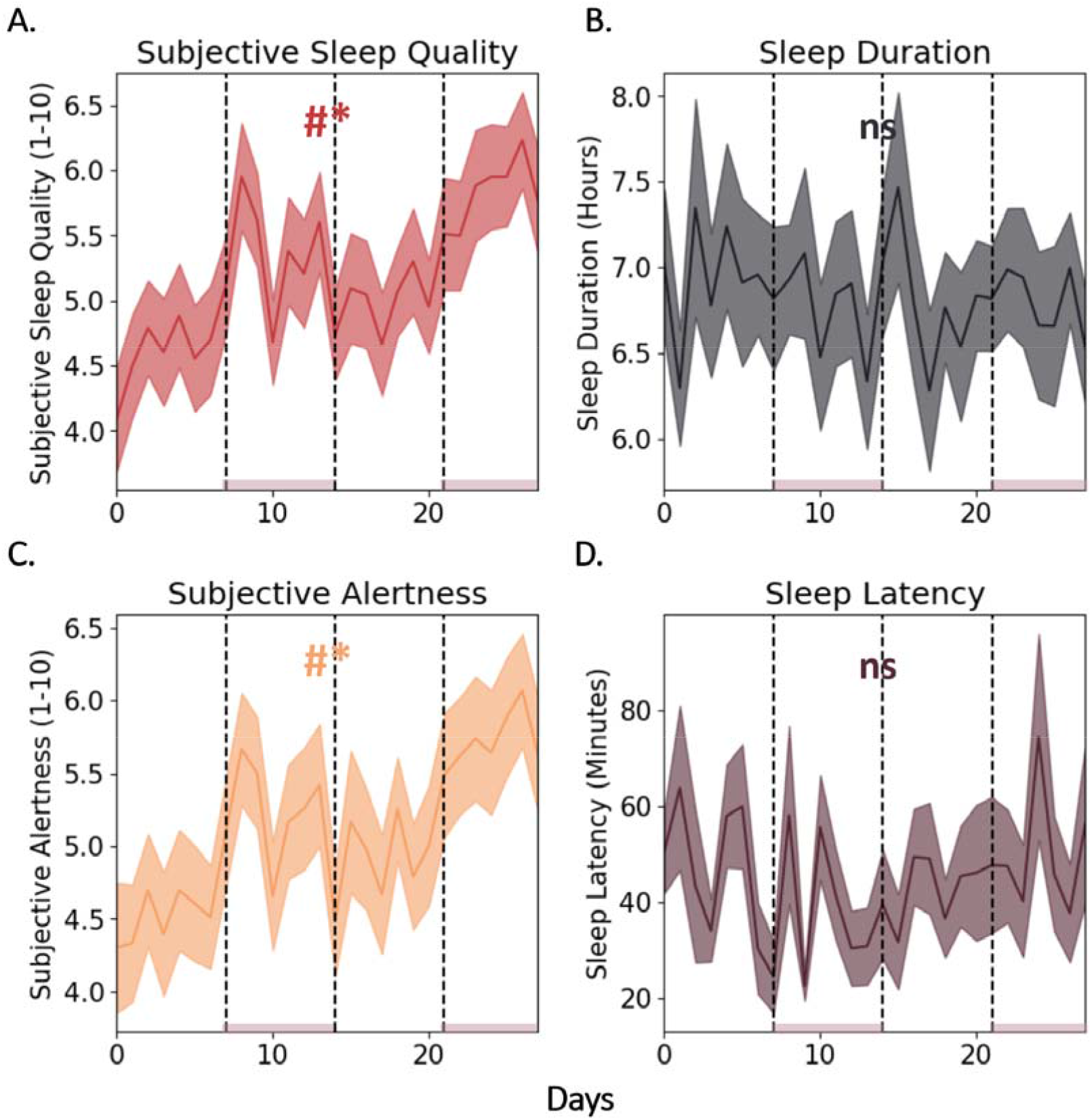
Daily Ratings of Subjective Sleep Quality and Alertness Trend Up Across Intervention. Subjective sleep quality, duration, subjective alertness, and sleep latency group means +/-SEM across the study. Shaded bars on the x-axis indicate weeks in which the sleep intervention was consumed. # indicates statistical Mann-Kendall trend over time across the entire study period. * Indicates statistical difference between weeks with and without the intervention.

## Discussion

Bedtime consumption of Sip2Sleep^**®**^, a sleep aid combining Montmorency tart cherry and Venetron^**®**^, increased subjective sleep quality and daytime alertness, reduced ISI, and reduced anxiety in a diverse cohort of adults with moderate to severe insomnia. We additionally observed that subjective sleep quality and daytime alertness trended upward across the study period; suggesting that these herbs may exert impacts on sleep on days subsequent to when they are taken, and that benefits may increase with longer use. ISI reduced on average from moderate to mild, whereas GAD-7 went from moderate to borderline mild-moderate.

These results are generally consistent with the limited reports of individual interventions with Montmorency tart cherry. Howatson et al.^28^, recently observed small but statistical elevations in melatonin in healthy volunteers who consumed Montmorency tart cherry concentrate as compared to a placebo juice, as well as increases in time in bed, sleep time, and sleep efficiency. Losso et al.,^30^ observed increases in PSG-measured sleep duration and sleep efficiency in a small cohort of older adults with insomnia, whereas a similar study of older insomniacs reported only reduction in nighttime wakeups^29^. More recently, Chung et al.^31^ observed that short term, higher-dose consumption of Montmorency tart cherry was associated with increases in time in bed, and decreases in night time wakeups and movement compared to a placebo in young female athletes.

Previous interventions with extracts of *apocynum venetum* produced similar results to those observed here. Recent reports indicate that *apocynum venetum* may modestly boost deep sleep time^33^, increase sleep quality, and decrease psychological stress^35^. Imafuku et al.^36^ similarly found that *apocynum venetum* leaf extract improved alertness upon waking and overall sleep quality (e.g., subjectively fewer nightly awakenings and easier falling asleep).

Previous studies are consistent in their reports of subjective sleep improvements, with some variability as to objective improvements. It is likely that errors in wearable sleep duration detection or staging obscure small but real changes in sleep structure across studies. Future studies relying on consistent high-quality sleep data, body temperature during sleep, or PSG may reveal a more consistent picture of how subjective sleep quality is related to physiological sleep with these interventions. Studies to date also highlight the need for larger placebo-controlled trials – potentially with longer washout periods and longer interventions. It is unknown if these effects would be greater in individuals with different severities of sleep disturbance, by sex, or how effects may vary by herbal dosage. Together, the combination of Montmorency tart cherry and Venetron^**®**^ consistently improves subjective metrics of sleep and anxiety in adults with moderate to severe insomnia.

Treatments involving herbal interventions, like those utilized in the present study, have some likely advantages over existing sleep medications^38,39^. Although apparently safe, over the counter melatonin has come under scrutiny due to wide variability in the actual melatonin content of tablets^40^. Moreover, there is a concerning rise in chronic insomnia combined with chronic prescription sleep aid use (e.g., Ambien/Zolpidem, Gabapentin) which may contribute to cognitive impairment and dementia^41–44^. These concerning effects highlight the importance for broader study and discussion of sleep aids with a long history of use, mild potency, and potentially better safety/tolerability.

## Conclusions and Future Directions

Evening consumption of Sip2Sleep® results in consistent improvement in subjective sleep quality and daytime alertness, and reduces insomnia and anxiety. These results are consistent with prior studies of individual interventions with either *prunus cerasus* or *apocynum venetum*. This is the first study, to our knowledge, to assess the synergistic effects of Montmorency Cherry and Venetron^**®**^ on sleep. Future placebo-controlled trials and longer-term interventions are necessary to determine the full impact of these extracts on sleep, and standardization of wearables used to assess sleep duration and latency may yield greater insight into how these compounds impact sleep structure.

## Methods

### Ethical Approval

This study and all procedures were approved by the Advarra Institutional Review Board (IRB) under Pro00068483. All participants gave informed consent. All research was performed in accordance with relevant guidelines and regulations. This protocol was registered at clinicaltrials.gov under NCT06299488, 07/03/2024.

### Recruitment and Inclusion Criteria

Participants were recruited via email outreach to People Science’s existing community, social media channels including Facebook, Instagram, LinkedIn and Reddit, outreach to patients of The Insomnia and Sleep Institute of Arizona (Gilbert and Scottsdale) and Sip2Sleep^®^ customer base. Pregnancy and lactation were exclusions due to the unknown risks of product use to fetuses and newborns. Individuals receiving Cognitive Behavioral Therapy for Insomnia (CBT-I) were excluded. Participants taking prescription hypnotics (e.g. zolpidem, zaleplon and benzodiazepines) or other classes of medication for sleep who were confirmed to be on a stable dose for at least 4 weeks were included. Participants taking over the counter or other products for sleep (e.g. melatonin, anticholinergics) or any cannabis products had to complete a 1 week wash out prior to the first day of sleep aid use and refrain from using these for the duration of the study.

Inclusion criteria consisted of moderate to severe insomnia was defined as in (Morin et al., 2011) as an ISI score > 15. Participants were also required to provide their own wearable device for estimation of sleep duration and sleep latency; and to provide informed consent to participate (including but not exclusive of inability to speak and read English, hearing or sight impaired, etc.).

### Study Design and Sleep Intervention

Sip2Sleep^®^ is a proprietary formula of Montmorency tart cherry extract and Venetron^**®**^, a purified, powdered extract derived from *Apocynum Venetum*. Recommended dose is ¼ teaspoon which contains 553 mg (1:5 fruit extract) of Montmorency tart cherry and 25mg of Venetron^®^ leaf extract. The compound was prepared as an oral dropper, consumed alone or in water 30-60 minutes prior to bedtime. The study was conducted over a period of four weeks, consisting of two weeks without intervention and two weeks with intervention as follows: (1 week off (baseline) - 1 week on - 1 week off - 1 week on) for each participant (**See Figure 1: Study Design**).

### Data Collection: Wearables and Questionnaires

During the four-week study, participants completed a daily morning questionnaire that asked about sleep quality (10-point sliding scale), daytime alertness (10-point sliding scale), and the time they laid in bed the night before (for calculation of sleep latency). Objective data for total sleep duration and bedtime latency were collected from participants’ personal wearable devices. Sleep latency was calculated as the difference between self-reported bedtime (time in bed with the lights out, attempting to sleep) and wearable-detected sleep onset. Participants also completed the Insomnia Severity Index (ISI) and Generalized Anxiety Disorder-7 Assessment (GAD-7) during baseline, at the beginning of each week, and at the end of the study.

### Data Management

Data were captured and organized using People Science’s HIPAA compliant platform, Chloe (<u>C</u>onsumer <u>H</u>ealth <u>L</u>earning and <u>O</u>rganizing <u>E</u>cosystem). The platform contains modules for building and managing forms / surveys, audited electronic consent forms, integration with wearable devices, data management and analytics using an integrated relational database. Data from completed assessments was automatically collected and de-identified prior to analysis.

### Data Analysis and Statistics

Data were analyzed in Python and R. Data were normally distributed and 2 tailed t-tests were used to evaluate differences between individual means in intervention and non-intervention conditions. Mann-Kendall trend over time tests were used to evaluate potential trends in daily questionnaire data across the study period. Averages were expressed as means +/-standard error of the mean.

## Data Availability

All data produced in the present study are available upon reasonable request to the authors

## Acknowledgements

The authors would like to thank S.W. for assistance with data organization and early analysis.

